# Benchmarking sex and gender incorporation into health and medical research, policy and education in Victoria prior to 2026: a mixed-methods study

**DOI:** 10.64898/2026.07.27.26358999

**Authors:** Sue Haupt, Tomer Parkiet, Inzela Mirza, Kyria Webster, Mila Waise, Zoe Wainer, Kim Kwan, Saraid Billiards, Kongchay Vongsaiya, Bronwyn Graham, Cara Tannenbaum, Lynn Riddell, Rachel Huxley, Séverine Lamon

## Abstract

**Objective:** To investigate and establish a baseline for sex and gender considerations in policy, research and curricula across the state of Victoria.

**Design and setting:** Victoria was selected as a case study for Australia, using a mixed-methods approach to examine health and medical university curricula, research organisation policies and research funding between 2020-2025 prior to mandated inclusion.

**Main outcome measures:** Primary outcomes include identification of predefined sex- and gender-related terms in university curricula descriptors and funded grant descriptions; and questionnaire responses from university course coordinators and organisational leads.

**Results:** Data mining across nine Victorian universities (318 courses/3383 units) identified ∼93% of units and ∼60% of healthcare courses lacked sex and gender terms in their descriptors. Among medical research organisations operating in Victoria, including peak bodies, research institutes, hospitals and universities, ∼70% (18/26) of the survey responders reported having no sex and gender policy. Rates of sex- and gender-term inclusion in research grants allocated in Victoria (3388) and Australia-wide (8974) validated Victoria as a case study for Australia for National Health and Medical Research Council (9.5%/8.9% respectively), Medical Research Future Funds (11%/10.2%), and Australian Research Council (4.8%/4.1%). One in nine Victorian awards from five government initiatives and one in ten from 12 non-government agencies included sex- and gender-term related terms in their guidelines.

**Conclusions:** These baseline metrics indicate that sex and gender are still not widely considered in the education and research ecosystems. These findings support the need to build inclusive research policy at a national and state level, and accreditation standards across university education.

## INTRODUCTION

Human health is influenced by biological sex characteristics (or “sex”, as defined in the SAGER guidelines [1]): usually male and female, and including chromosomes, gene expression, hormone function, etc; together with socially constructed gender identities (or “gender”, as defined in the SAGER guidelines [1]), including behaviours, social roles, etc [2–4]. Ignoring these factors creates inequity in health care [5]. Despite clear evidence of the benefits in having sex and gender inclusive policies on health outcomes, Australia lags over a decade behind North America [6] and Europe in this area [7, 8]. These gaps have important health consequences: sex and gender influence disease risk, presentation and treatment response, yet women’s health remains inadequately researched, while trans and gender-diverse people and people with innate variations of sex characteristics remain poorly represented in health research and education despite experiencing substantial health disparities [9, 10]. Given this strong evidence base, failure to consider these important health determinants is a growing concern (2) and has prompted reforms.

Statements advocating practice changes were published by the Australian Association of Medical Research Institutes (AAMRI) in 2023, in a grass roots bid for self-determination [8]; followed in 2024 by policy guidance from the National Health and Medical Research Council (NHMRC) through its *Statement on Sex, Gender, Variations of Sex Characteristics and Sexual Orientation in Health and Medical Research* [11]. A landmark requirement was then issued by NHMRC in November 2025 [12] stating that grant submissions from the beginning of 2026 must: *consider sex, gender, variations of sex characteristics or sexual orientation in every stage of their research project; and describe how they have done this in their applications for funding* [11], ensuring that inclusive research design extends beyond participant representation to ensure that sex, gender and related diversity are appropriately considered across the research lifecycle, from study design and recruitment through to data collection, analysis, reporting and interpretation. This statement signals progress at the national level, yet its implications for implementation and alignment across the wider research and funding landscape remain uncertain. In addition, no equivalent mandate currently exists for the integration of these considerations within health and medical education curricula in Australia.

While a few focused studies before 2026 offered early insights into sex and gender exclusion from various aspects of the health and medical sector [7, 13–15], comprehensive measurements are lacking across Australian health and medical education and research. Victoria hosts the largest concentration of health and medical research funding and infrastructure in Australia, including near 20 independent medical research institutes (MRIs) and two globally leading universities for health and medical professions (Melbourne and Monash Universities). Victoria attracts ∼40% of NHMRC funding [16], despite its disproportionately smaller population size [17], making it both a national leader and a relevant lens through which broader Australian research and policy trends can be examined. Our aim was to score a baseline at time zero, anchored on the five years preceding the 2026 pivotal NHMRC compliance requirement, to enable future assessment of the policy impact. Using Victoria as a case study for Australia, we document the extent of sex and gender integration into university curricula, healthcare policies governing health and medical research and research funding policy and awards. The development and validation of an Artificial Intelligence (AI) tool served this primary aim by providing a scalable method for measuring the baseline. Finally, we proposed a potential pathway for translating the evidence into future policy and practice.

## METHODS

### Ethical approval

Ethics approval was granted by [Administrating Institution] University Human Ethics Committee (2024/HE000736).

### Study design

Cross-sectional data were collected either from targeted surveys or by data mining of publicly available information using both manual and automated methods for output accuracy and efficiency (**Figure 1**).

**Figure 1.**
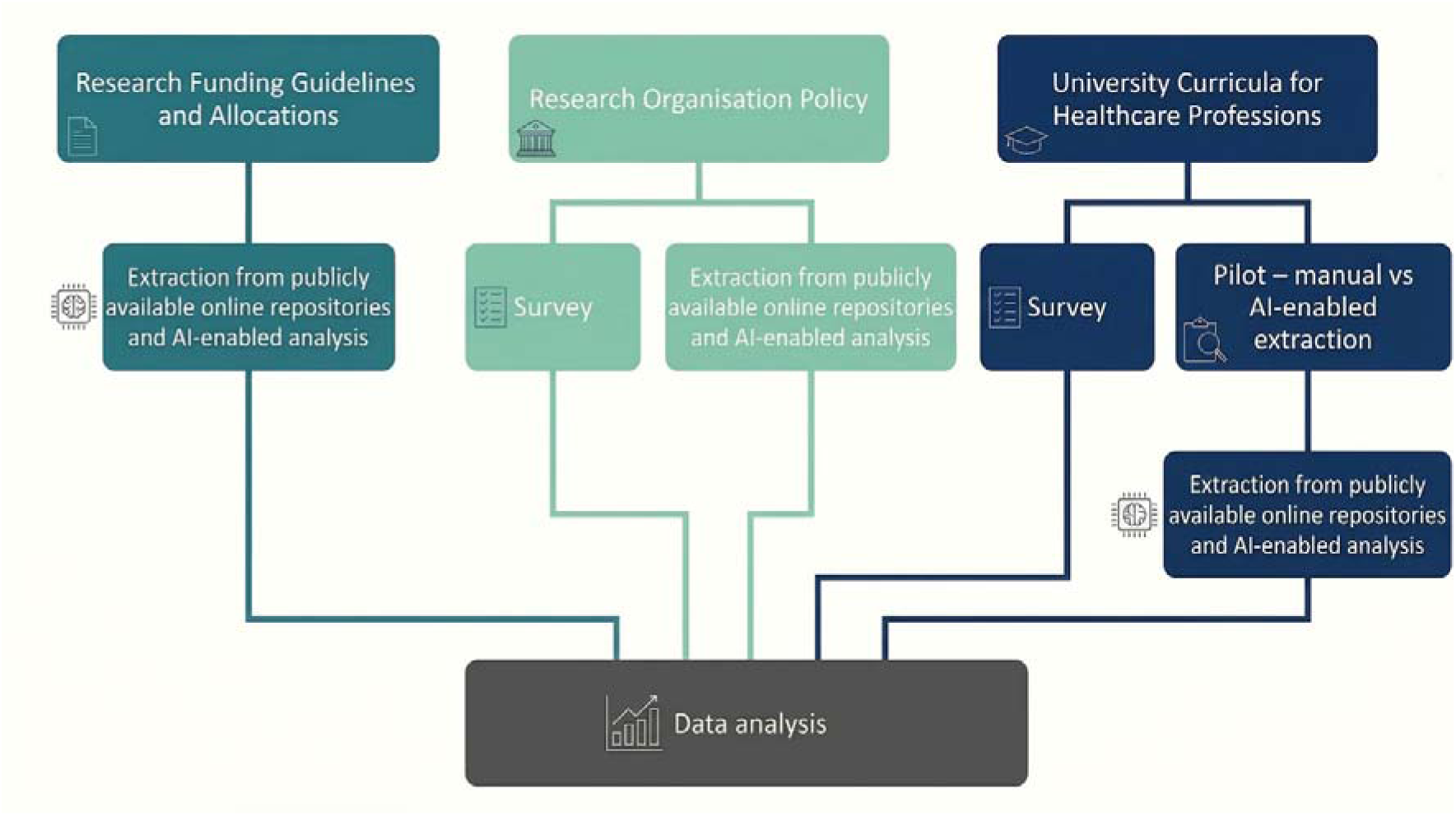
Analytical workflow of the study. AI = Artificial Intelligence.

### University Curricula for Healthcare Professions

Eligible university courses (defined as a complete program of study leading to a university qualification) included: medicine, dentistry, nursing and the 27 allied health professions that are recognised by the Victorian Government [13] comprising bachelor’s, master’s degrees and professional doctorates. Australian Health Practitioner Regulation Agency (AHPRA) accredited courses were prioritised.

We first conducted a pilot study assessing sex and gender term inclusion in Deakin University’s Faculty of Health curricula to compare the efficacy and accuracy of survey-based, manual and automated data-mining approaches and inform scale-up across Victorian universities.

### Survey-based validation of the data mining approach for university curricula

A Qualtrics questionnaire (Qualtrics, Provo, UT, United States of America [USA]; https://www.qualtrics.com) was developed to identify sex and gender topics in university course content (**Supporting Information S1.1**). The survey conformed to Checklist for Reporting Results of Internet E-Surveys (CHERRIES [18]) **(Supporting Information S2)** and was tested across the 35 relevant courses within Deakin University’s Faculty of Health **(Supporting Information S3).** Response rate was 91% (32/35). Reference to sex and/or gender was reported in 78% of these courses, although each course is comprised of many units (defined as an individual subject or component undertaken within a course) and mention of sex/gender-terms in a single unit was sufficient to qualify the entire course for inclusion. Of these courses, 72% of references were in a non-reproductive context and 38% in a reproductive context, where some courses taught units referencing both. Deakin University course coordinators widely considered (72%) that a directive to guide the inclusion of sex and gender into curricula would be favourably received. This survey approach identified course-level detail without gathering comprehensive information regarding individual unit constituents. Dissemination of the questionnaire to individual unit coordinators across nine universities was considered operationally unfeasible, leading to the initiation of a data-mining strategy.

### Data mining

Publicly available online repositories hosted by university websites allowed access to relevant course and unit outlines from Victorian universities. Course descriptors and unit guides were accessed for the 2024-2025 period. Courses within the study scope were first identified, after which the associated datasets were retrieved through investigator-led extraction (“manual”). A model custom Python-based web scraper (Python v3.11) (“automated”) was developed to evaluate scalability and provide a proof of principle (full code available at: https://github.com/severinelamon-prof/Sex-and-gender-incorporation-in-research-in-Victoria/tree/main) (**Figure 2**, upper tier).

**Figure 2.**
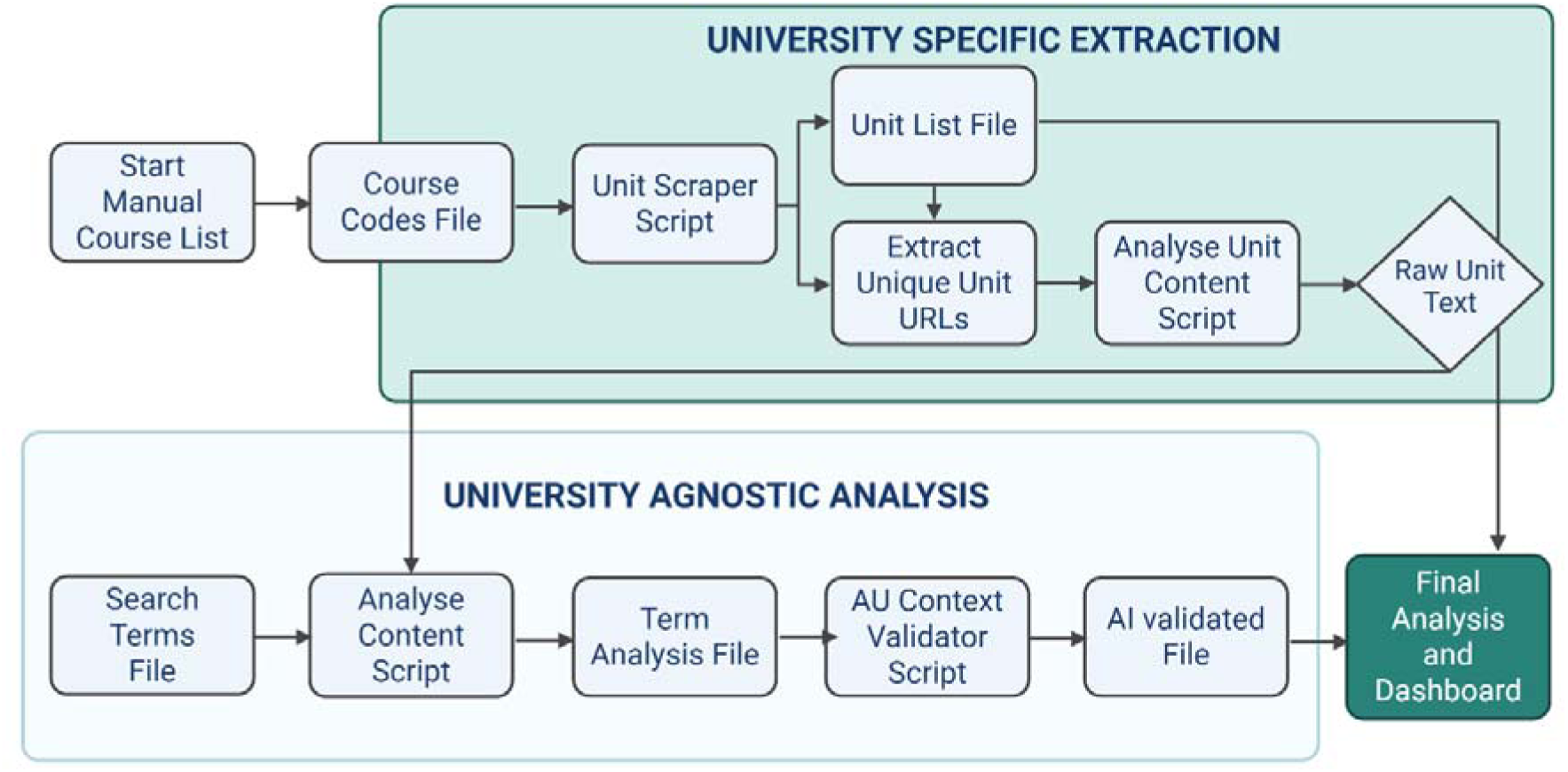
Flow Diagram of the Informatics pipeline for mining and analysing university curricula. The four phases were (1) University-specific data extraction (a Python-based web scraper); (2) University-Agnostic content analysis; (3) Context extraction; and (4) AI-Driven contextual validation that filtered for substantive relevance, relying on Google Gemini Application Programming Interface for nuanced classification. Image was created in https://BioRender.com.

### Analytical strategies

Key search terms for sex and gender (**Table 1**) were compiled through consultation with topic experts. Term searches were undertaken manually with Microsoft software (Microsoft Corporation, Redmond, WA, USA). Manually searched term matches were validated using AI language models to interrogate the extracted unit datasets. These searches were prompted either in Claude Sonnet 4.5 (Anthropic, San Francisco, CA) or ChatGPT Pro (Open AI). In practice, a manual feedforward loop between manual and these automated pipelines was adopted to optimise outcomes. Throughout the study, the presence of sex- and gender-related key terms was used as the measure of incorporation of sex and gender considerations across these different contexts. In addition, we created a purpose-built, automated term matching computational tool (using Base44, Inc., San Francisco Bay Area, California, USA) (**Supporting Information S4** and https://github.com/severinelamon-prof/Sex-and-gender-incorporation-in-research-in-Victoria/tree/main). This was applied to the uploaded documents generated with our Python-coded web-scraper (**Figure 2**, lower tier). Results were summarized as the proportion of unit or course outlines that tested positive for sex and gender inclusion terms. Deakin University course handbook analyses closely recapitulated the survey data (77%), validating the pipeline with the added value of enabling comprehensive unit level analyses.

**Table 1.**
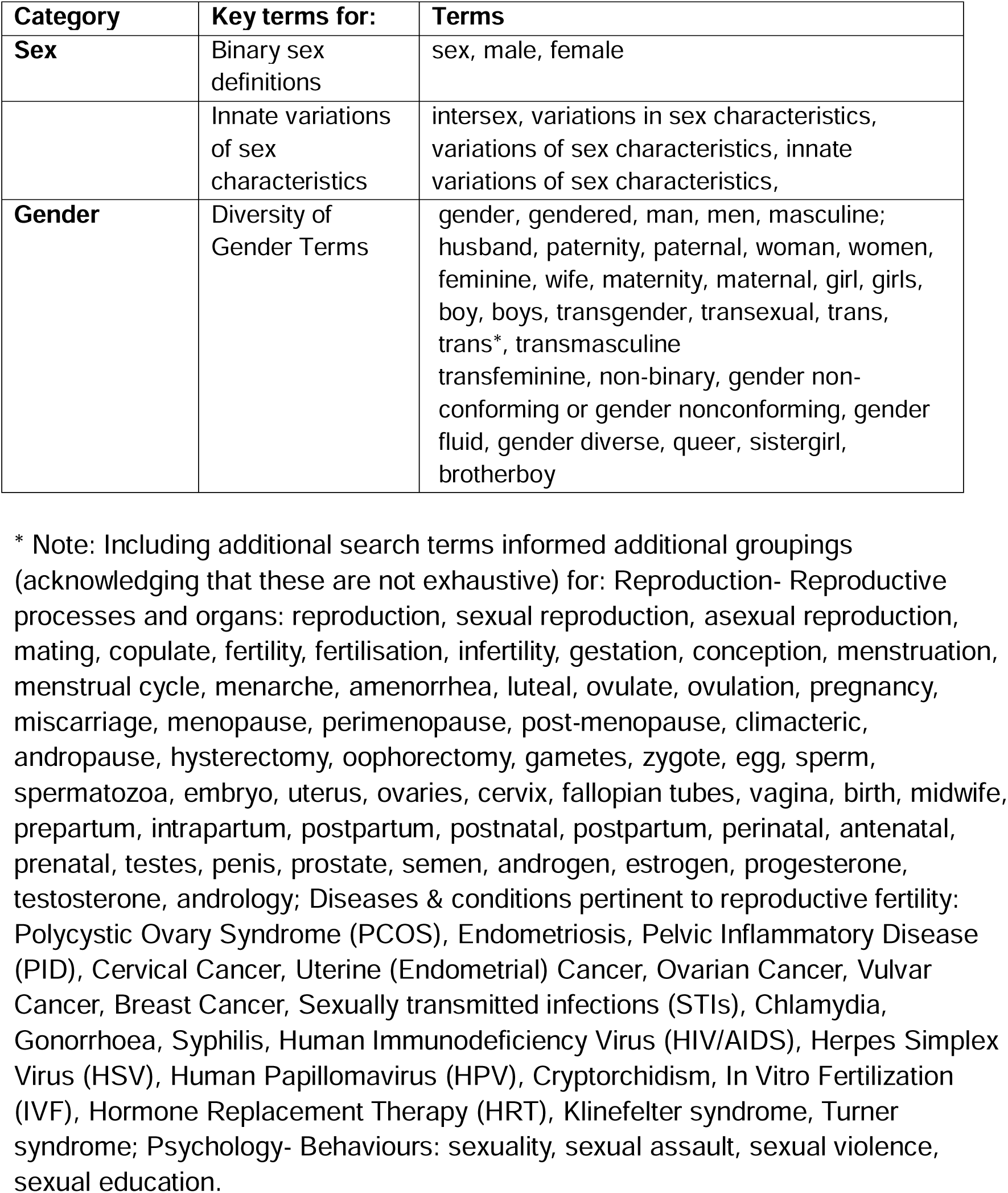
– Key search terms.

### Research organisation policy

#### Research organisation policy survey

A Qualtrics questionnaire was developed to identify sex and gender-inclusive research policies within Victorian health and medical research organisations (**Supporting Information S1.2**). The survey conformed to CHERRIES [18]. Research organisation leads at government-funded Victorian universities were approached to complete the relevant survey assessing the extent of sex and gender inclusion within their organisation. Information was gathered on existing relevant policies or directives, publicly available sources and organisational interest in introducing such policies where they did not previously exist.

#### Data mining and analytical strategies

Publicly available organisation websites enabled access to policy documents from Victorian health and medical research organisations. Online organisation profiles were analysed for policies guiding sex and gender inclusive research as described above, and results summarized as the proportion of policies, guidelines and/or directives that tested positive for sex and gender inclusion terms.

### Research funding guidelines and allocations

#### Data mining and analytical strategies

Publicly available online repositories enabled access to funded application summaries. Funded research scheme descriptors and guidelines were identified online and downloaded for analysis. Analyses were conducted as described above, and results summarized as the proportion of funded grants or guidelines that tested positive for sex and gender inclusion terms.

## RESULTS

### University Curricula for Healthcare Professions

The scalability of the data-mining approach enabled analysis across nine publicly funded universities, encompassing 318 courses and 3,383 unique units (**Figure 3**).

**Figure 3.**
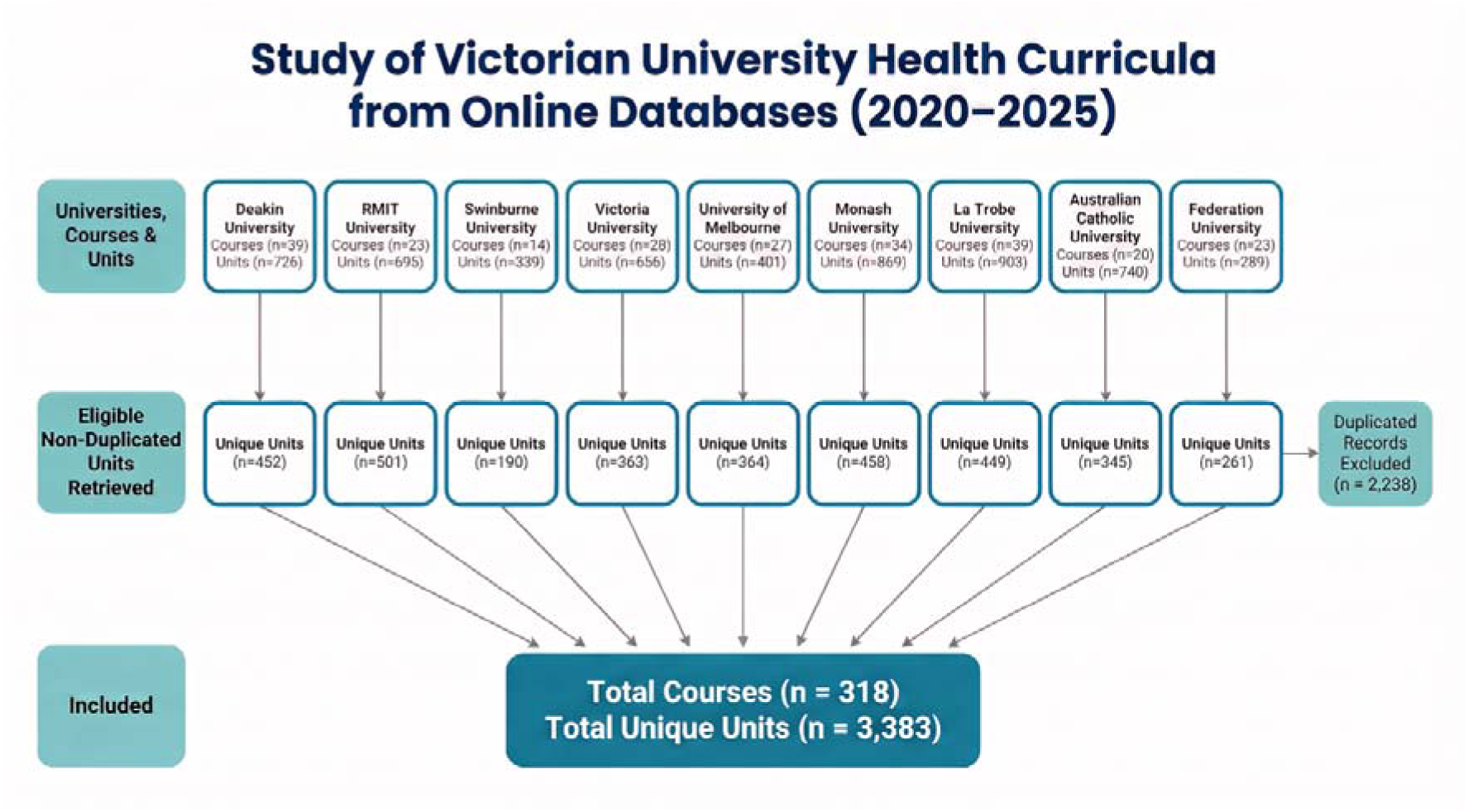
Identification and inclusion of Victorian university health curricula from publicly available online databases (2020–2025). Health-related course and unit information was retrieved from the publicly available online databases of nine Victorian universities: Deakin University (39 courses; 729 units), RMIT University (23 courses; 695 units), Swinburne University (14 courses; 339 units), Victoria University (28 courses; 656 units), University of Melbourne (27 courses; 401 units), Monash University (34 courses; 689 units), La Trobe University (39 courses; 903 units), Australian Catholic University (20 courses; 740 units), and Federation University (23 courses; 289 units). Following removal of units duplicated within or across eligible courses at each institution, 452 unique units were retained from Deakin University, 501 from RMIT University, 190 from Swinburne University, 363 from Victoria University, 364 from the University of Melbourne, 458 from Monash University, 449 from La Trobe University, 345 from Australian Catholic University, and 261 from Federation University. Overall, 318 courses comprising 3,383 unique units were included in the final curriculum analysis. A total of 2,238 duplicated unit records were excluded prior to analysis. RMIT = Royal Melbourne Institute of Technology. Image was generated using Nano Banana, a multimodal image-generation model developed by OpenAI (San Francisco, CA, USA), accessed February 2026.

Across the nine publicly funded Victorian Universities, ∼7% (244/3383, range <1-19%) of all the unique 3383 teaching units reference sex or gender terms in unit outlines within the 318 courses training health professionals. Overall ∼40% (127/318, range 9-81%) of all courses teaching health professions at nine Victorian Universities had at least one unit outline referencing sex or gender terms **(Supporting Information S5).**

### Research organisation policy

Survey response rate was 74% (26/35) for Victorian medical research organisations, including peak bodies (3/4), Association of Australian Medical Research Institutes (AAMRI) affiliated MRIs (16/16), Victorian publicly funded universities (3/10) and Victorian hospitals undertaking research (4/5). The survey response rate was 100% (16/16) amongst AAMRI affiliated MRIs (**Table 2**).

**Table 2.**
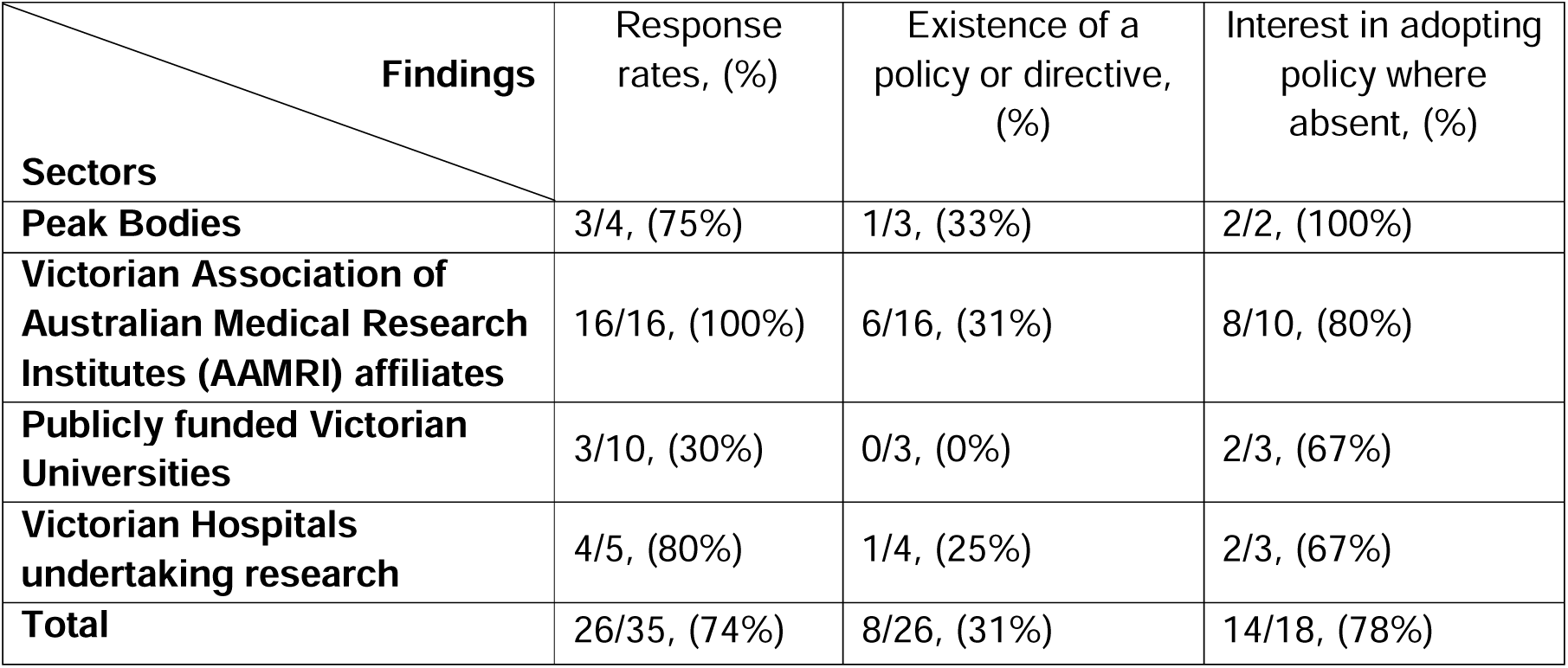
– Research policy survey outcomes in Victorian research organisations.

| Findings<br>Sectors | Response rates, (%) | Existence of a policy or directive, (%) | Interest in adopting policy where absent, (%) |
| --- | --- | --- | --- |
| **Peak Bodies** | 3/4, (75%) | 1/3, (33%) | 2/2, (100%) |
| **Victorian Association of Australian Medical Research Institutes (AAMRI) affiliates** | 16/16, (100%) | 6/16, (31%) | 8/10, (80%) |
| **Publicly funded Victorian Universities** | 3/10, (30%) | 0/3, (0%) | 2/3, (67%) |
| **Victorian Hospitals undertaking research** | 4/5, (80%) | 1/4, (25%) | 2/3, (67%) |
| **Total** | 26/35, (74%) | 8/26, (31%) | 14/18, (78%) |

Among survey respondents, ∼70% (18/26) lacked sex and gender directive or policy framework (**Table 3, Supporting information S6.1 and S6.2**). In comparison, AI analyses reported directives absent from ∼80% of organisations (28/35) (**Supporting information S6.3**); with only AAMRI identified to have policy. This discrepancy suggests that not all directives are publicly available.

**Table 3.**
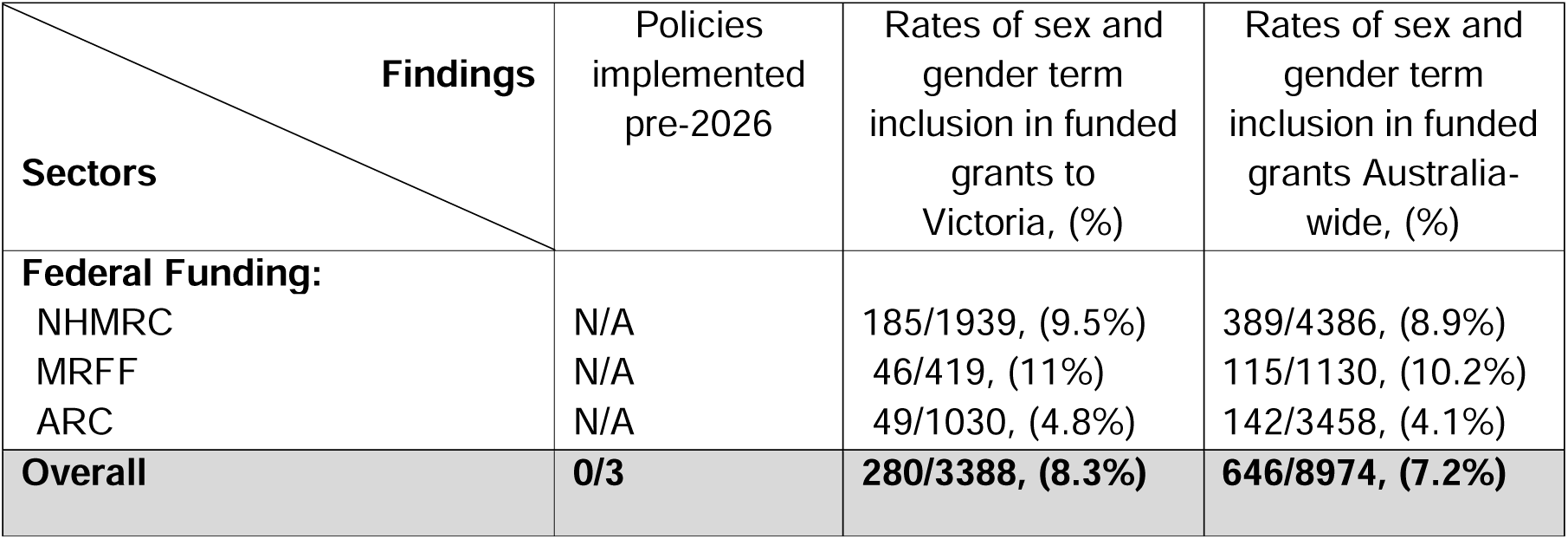
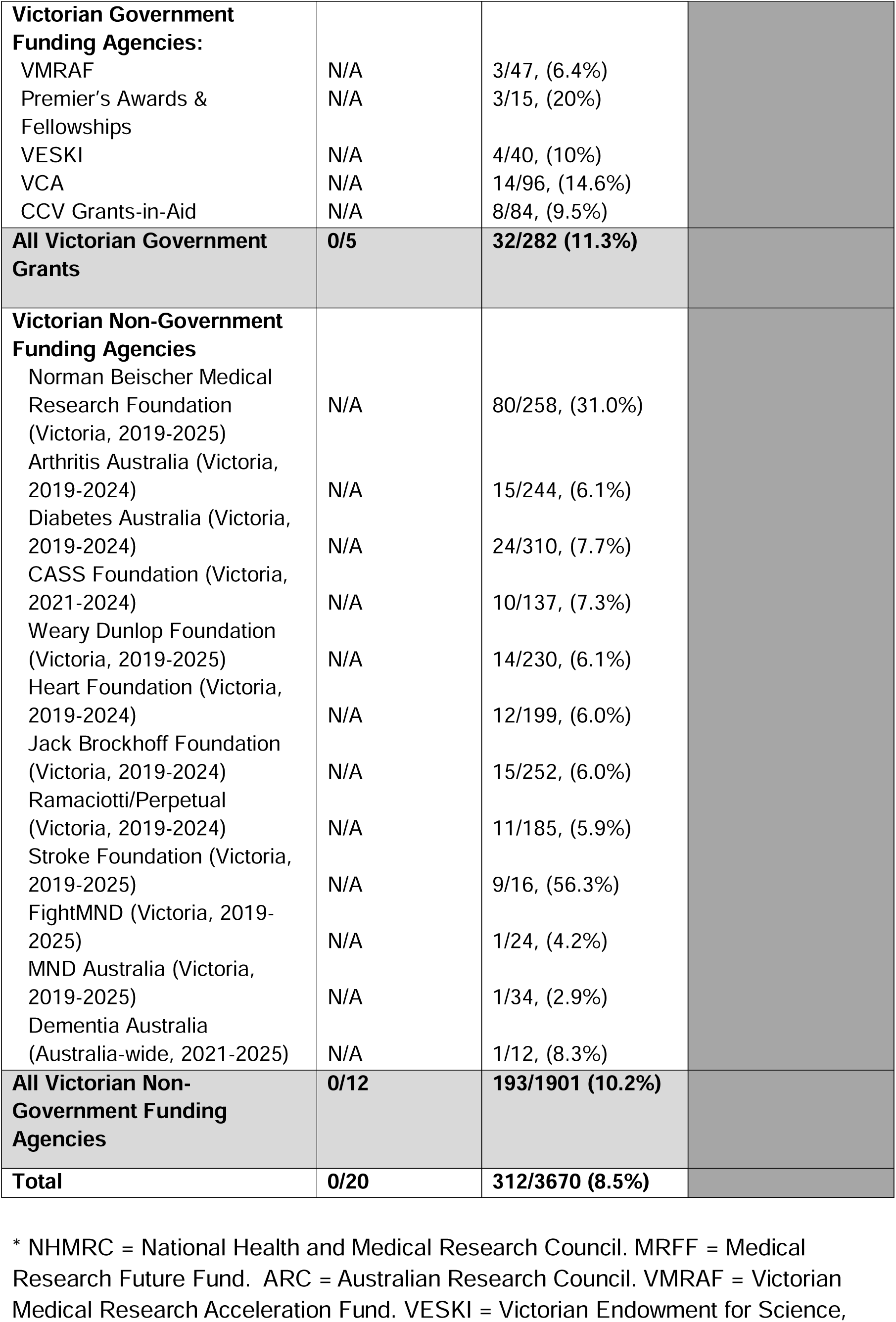
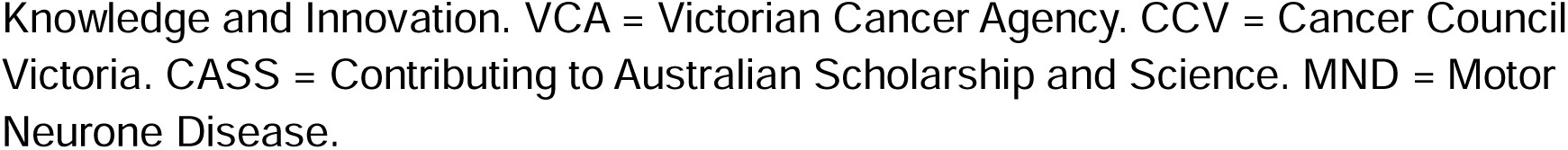
– Sex and gender policy and inclusions rates in federal and Victorian funding schemes 2020-2025.

| Findings<br>Sectors | Policies implemented pre-2026 | Rates of sex and gender term inclusion in funded grants to Victoria, (%) | Rates of sex and gender term inclusion in funded grants Australia-wide, (%) |
| --- | --- | --- | --- |
| **Federal Funding:** |  |  |  |
| NHMRC | N/A | 185/1939, (9.5%) | 389/4386, (8.9%) |
| MRFF | N/A | 46/419, (11%) | 115/1130, (10.2%) |
| ARC | N/A | 49/1030, (4.8%) | 142/3458, (4.1%) |
| **Overall** | **0/3** | **280/3388, (8.3%)** | **646/8974, (7.2%)** |
| **Victorian Government Funding Agencies:** |  |  |  |
| VMRAF | N/A | 3/47, (6.4%) |  |
| Premier's Awards & Fellowships | N/A | 3/15, (20%) |  |
| VESKI | N/A | 4/40, (10%) |  |
| VCA | N/A | 14/96, (14.6%) |  |
| CCV Grants-in-Aid | N/A | 8/84, (9.5%) |  |
| **All Victorian Government Grants** | **0/5** | **32/282 (11.3%)** |  |
| **Victorian Non-Government Funding Agencies** |  |  |  |
| Norman Beischer Medical Research Foundation (Victoria, 2019-2025) | N/A | 80/258, (31.0%) |  |
| Arthritis Australia (Victoria, 2019-2024) | N/A | 15/244, (6.1%) |  |
| Diabetes Australia (Victoria, 2019-2024) | N/A | 24/310, (7.7%) |  |
| CASS Foundation (Victoria, 2021-2024) | N/A | 10/137, (7.3%) |  |
| Weary Dunlop Foundation (Victoria, 2019-2025) | N/A | 14/230, (6.1%) |  |
| Heart Foundation (Victoria, 2019-2024) | N/A | 12/199, (6.0%) |  |
| Jack Brockhoff Foundation (Victoria, 2019-2024) | N/A | 15/252, (6.0%) |  |
| Ramaciotti/Perpetual (Victoria, 2019-2024) | N/A | 11/185, (5.9%) |  |
| Stroke Foundation (Victoria, 2019-2025) | N/A | 9/16, (56.3%) |  |
| FightMND (Victoria, 2019-2025) | N/A | 1/24, (4.2%) |  |
| MND Australia (Victoria, 2019-2025) | N/A | 1/34, (2.9%) |  |
| Dementia Australia (Australia-wide, 2021-2025) | N/A | 1/12, (8.3%) |  |
| **All Victorian Non-Government Funding Agencies** | **0/12** | **193/1901 (10.2%)** |  |
| **Total** | **0/20** | **312/3670 (8.5%)** |  |

\* NHMRC = National Health and Medical Research Council. MRFF = Medical Research Future Fund. ARC = Australian Research Council. VMRAF = Victorian Medical Research Acceleration Fund. VESKI = Victorian Endowment for Science,
Knowledge and Innovation. VCA = Victorian Cancer Agency. CCV = Cancer Council Victoria. CASS = Contributing to Australian Scholarship and Science. MND = Motor Neurone Disease.

Six MRIs reported a directive or policy (**Table 2**, **Supporting information S6.3**). Two thirds (10/16) were unaware of a directive or policy from within their MRI; and 80% (8/10) of these MRIs were interested in adopting one. Survey response rate was 80% (4/5) amongst Victorian hospitals. A single hospital (1/4, 25%) had directives or policy in place. Two hospitals expressed an interest in introducing a policy.

Overall, from the ∼70% (18/26) of organisations undertaking health and medical research in Victoria lacking directives to guide inclusion of sex and gender in research in Victoria, 78% (14/18) expressed interest to adopt policy.

### Research funding guidelines and allocations

Across 2020 to 2025, there were low numbers of federally funded, health and medicine grants that included sex and gender terms (term inclusive grant descriptors/ all grants) in Victoria (280/3388) and Australia-wide (646/8974) for the three major funders: NHMRC (185/1939, 9.5%/ 389/4386, 8.9%), Medical Research Future Fund (MRFF; 46/419, 11%/ 115/1130, 10.2%) and Australian Research Council (ARC; 49/1030, 4.8%/ 142/3458, 4.1%), validating Victoria as a case study for Australia (**Table 3**; **Supporting Information S7.1)**.

Low rates of term inclusion were also measured among five Victorian government funding agencies: 11.3% (32/282) of grants (excluding specific funding for women’s health e.g. Catalyst grants) and 12 non-government agencies: 10.2% (193/1901) of grants (**Supporting Information S7.2**). Explicit policies mandating sex and gender inclusive research were not identified by data mining to be in place prior to 2026 (20/20 schemes; **Table 3**), although partial policy was reported for some organisations (7, 15). NHMRC and MRFF are the first to define obligatory requirements to be implemented from 2026 (**Supporting Information S7.3**). **Figure 4** provides a visual summary of the funding (Figure 4A), policy (Figure 4B) and curriculum (Figure 4C) components of the study.

**Figure 4.**
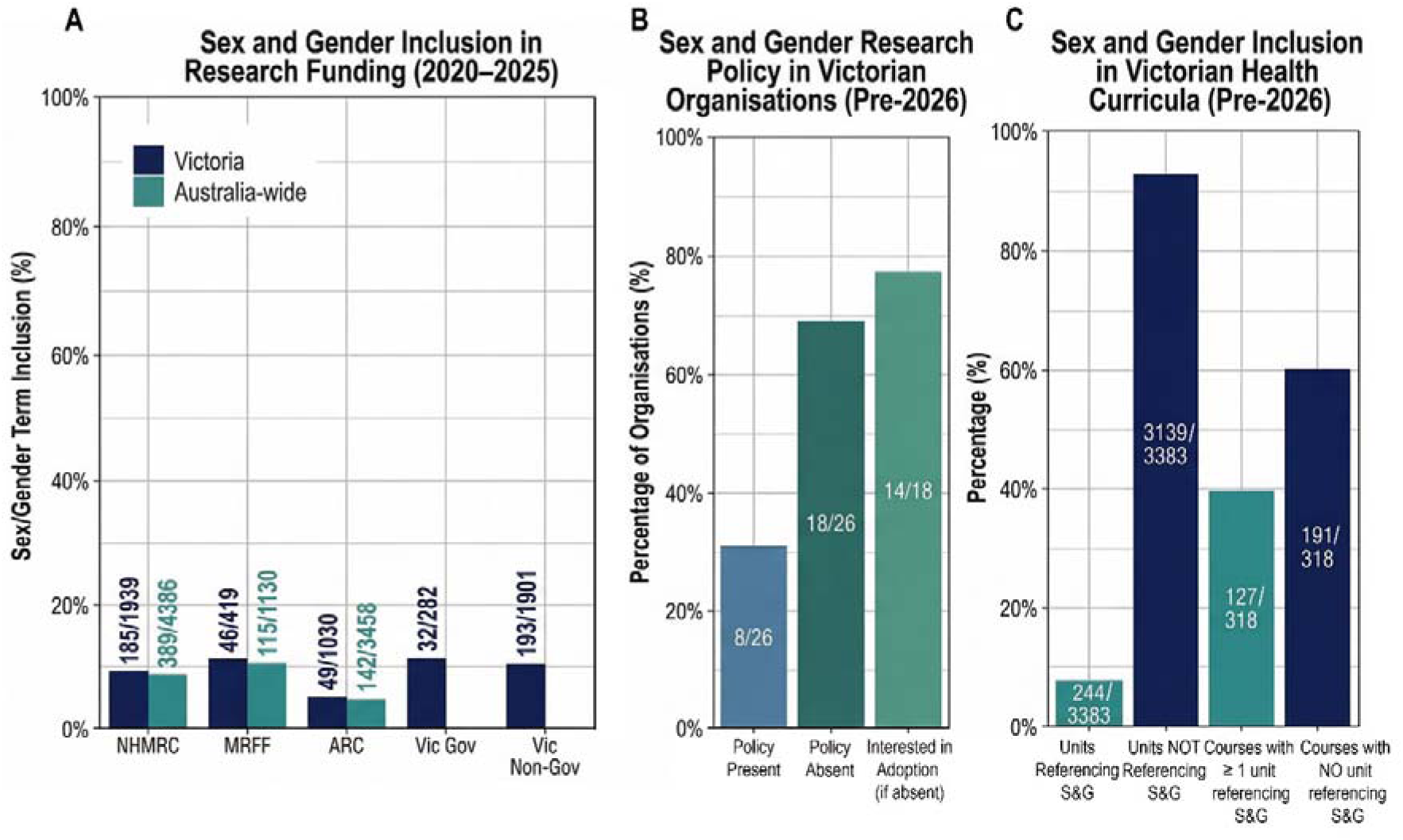
Visual summary of the study results. A) Sex and Gender Inclusion in Research Funding. B) Sex and Gender Inclusion in Victorian Research Organisation. C) Sex and Gender Inclusion in Victorian Health Curricula. S&G = Sex and Gender. NHMRC = National Health and Medical Research Council. MRFF = Medical Research Future Fund. ARC = Australian Research Council. VIC Gov = Victorian Government. Image was generated using Nano Banana, a multimodal image generation model developed by OpenAI (San Francisco, CA, USA), accessed February 2026.

## DISCUSSION

Without gauging the extent to which sex and gender are referenced as health determinants at a baseline timepoint, it will be impossible to measure whether new mandates and other future initiatives result in change. To generate an instrument suited to contemporary and prospective use, we constructed an informatic barometer. We applied this, in advance of the 2026 NHMRC compliance requirements [12], to demarcate a sharp pre-implementation benchmark across the Victorian education and research facets of the health sector.

Sex- and gender-related terms were absent from university policy documents, 93% of unit guides, and 60% of courses training health professionals across nine public universities in Victoria. Over two-thirds of medical institutions conducting research in Victoria lacked sex and gender directive of policy framework, and about 10% of all surveyed grants included a sex or gender component. These data establish a useful baseline against which to evaluate the impact of the forthcoming NHMRC compliance requirements. They also highlight policy gaps in health and medical education, where no equivalent mandate exists, despite individual Victorian universities taking the lead [19]. Because the evidence generated through research ultimately informs clinical guidelines, and the training of health professionals determines how this evidence is interpreted and applied in health and medical services, aligning expectations across research and professional curricula is critical. Trans and gender-diverse people and people with innate variations of sex characteristics also remain inadequately represented in health and medical research and curricula, limiting the evidence and workforce knowledge needed to address their specific health needs and persistent health disparities [10, 20, 21]. Strengthening both domains will result in improved health and medicine by ensuring that sex- and gender-informed evidence is both generated and effectively translated into clinical practice. This is relevant to health equity, particularly for women.

Our systematic documentation of pervasive gaps in sex- and gender-considerations in Australian research funding, research policy and education within Victoria aligns with the maxim that education underpins knowledge. This is complemented by earlier, more focused studies by others that identified: the absence of women’s health topics in Australian medical school curricula and texts across 2023-2024 [14]; no dedicated men’s health courses or electives in Australian university health curricula in 2022-2023 [22]; a lack of consistent, dedicated directives across a diverse group of Australian research organisations in 2022 [13]; and no standardised incorporation of sex and gender considerations in clinical practice guidelines within Australia [15]. In line with this, we identified that sex and gender term inclusion rates in National and State research funding applications did not exceed 12% overall. The low levels of consideration of these fundamental determinants of health in funding applications reflect the absence of obligatory standards from funding bodies and a lack of directives from Victorian research organisations.

### Strengths and limitations

Our multi-methods approach offers insight into the worth of manual and automated analyses for performing large scale, multi-sector analyses. Our university findings were sourced entirely from available handbooks, which are the public face of the university curricula and do not include comprehensive course content, assessment tasks and other learning materials. It is therefore possible some of the units and courses surveyed did have a sex and gender component, although it was not featured in the unit guide or course outline, respectively. However, the close concordance observed between survey responses and data extraction results in the pilot study suggests that unit guides and course outlines provide a reasonable proxy for the broader curriculum. AI capacity to mine sourced data significantly enhances the generation of accurate analyses at a fraction of the cost and resources of manual analyses. The accuracy of the output, however, relies on the precision of the prompt, consistency of the AI to precisely repeat an exact function, and capacity to identify errors and correct accordingly.

Our online data mining approach, solely based on publicly available content, required no institute consent or ethical approval, and was devoid of stakeholder engagement. In contrast, our questionnaires involved direct communication with university stakeholders on the chosen topic, with the advantage of probing in-depth insights from the participants, and high-level organisational authorities engaged to provide consent.

Finally, our assessment captured the explicit integration of sex and gender rather than gender mainstreaming more broadly; implicit practices, such as attention to gender representation in teaching materials or the selection of researchers and case studies, may therefore not have been captured by our approach.

### Lessons beyond this immediate study

The low representation of sex- and gender-related terms in unit guides and course outlines for health professional training, as well as in research organisation policies, research awards and funding guidelines suggests that sex and gender considerations remain poorly integrated across research funding, organisational policy, and health professional education in Victoria. These glaring inadequacies have stark implications for research quality, clinical training, and health equity. Specifically, the deficiencies exposed in teaching curricula risk feeding inadequately prepared workers into the healthcare system beyond medical courses alone [14], and are likely to be a major contributor to the reported inequity in healthcare access and health outcomes among women. In **Table 4**, we outline decisive initiatives that responsible authorities may action to reform Australian health practices. These recommendations were developed in collaboration with lead authors of the final report and in consultation with the Victorian Department of Health. The recommendations are informed by the study findings and the current Australian context, including existing resources, governance frameworks and academic, policy, and research infrastructure in place. The recommendations aim to improve the inclusion of sex and gender in the practice of psychology [24] and represent context-informed, consensus-based recommendations rather than interventions directly evaluated in this study. To improve the incorporation of sex and gender considerations, international experience demonstrates that progress can be supported through a combination of policy requirements, incentives, researcher education and resources, integration into research assessment and peer review, and mechanisms for monitoring and accountability. In the Australian context, these approaches could be guided by existing theory-of-change frameworks that emphasise coordinated action across research, policy and practice, and adapted to the governance structures, resources and responsibilities of individual organisations [13, 25, 26].

**Table 4.**
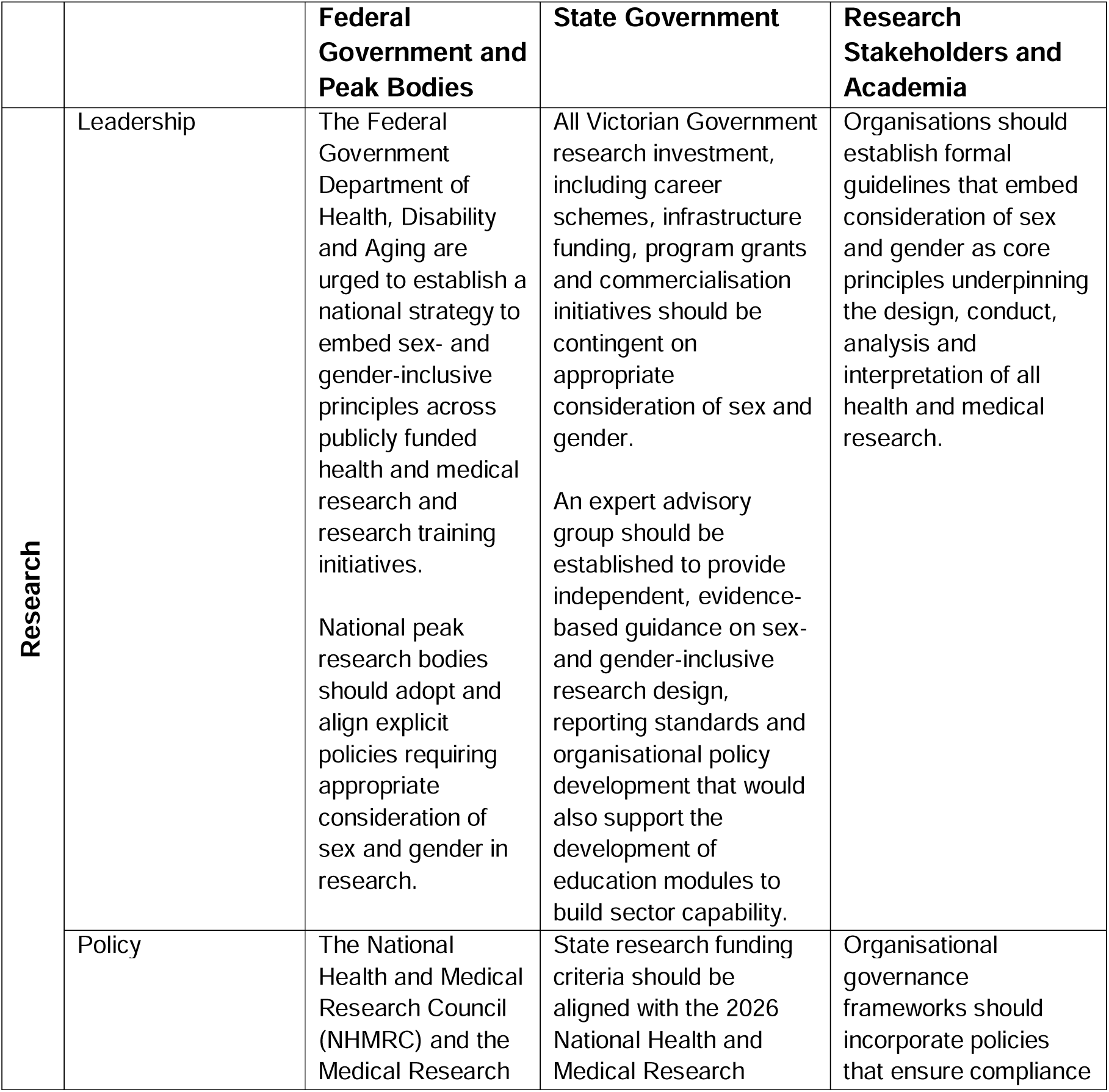
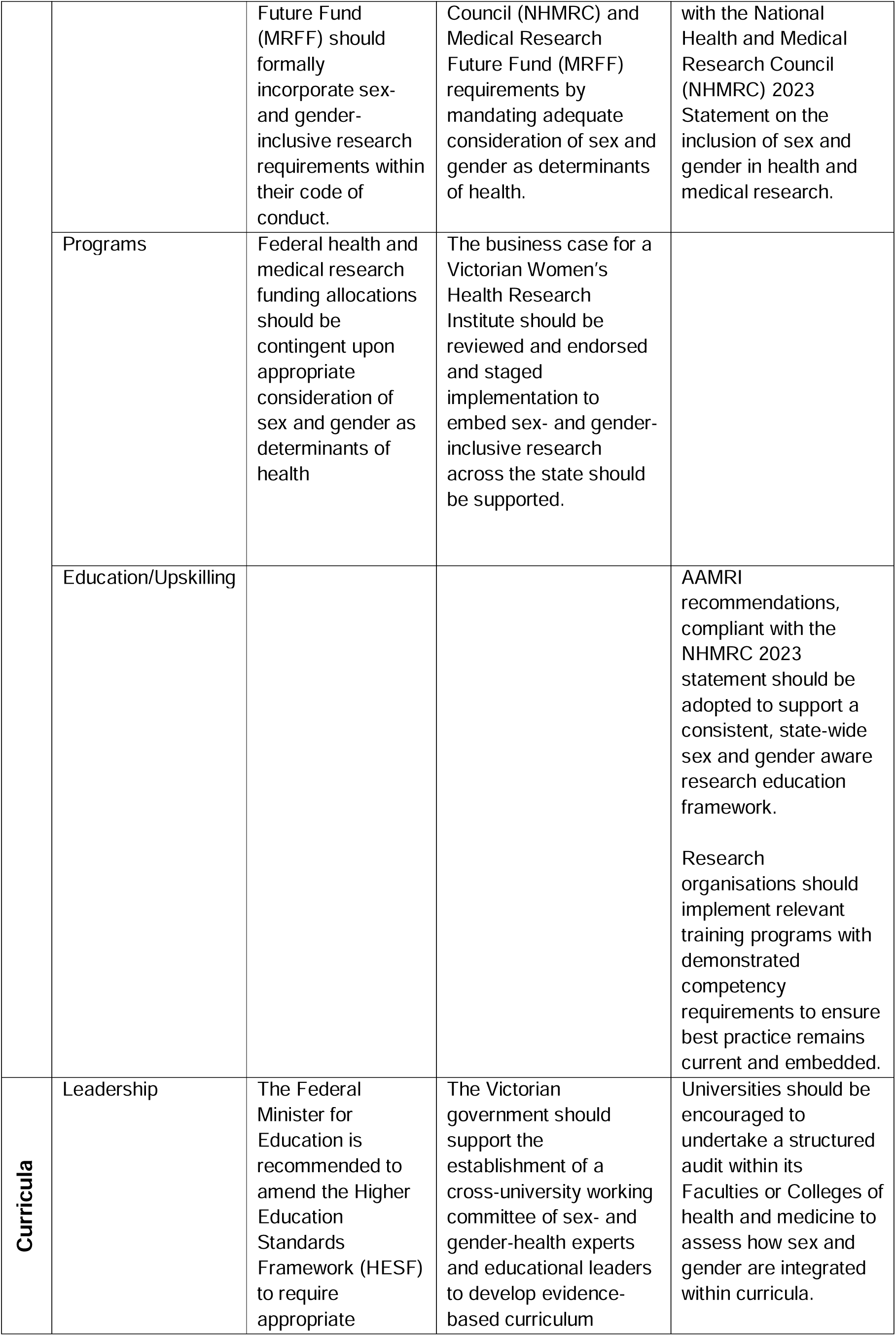
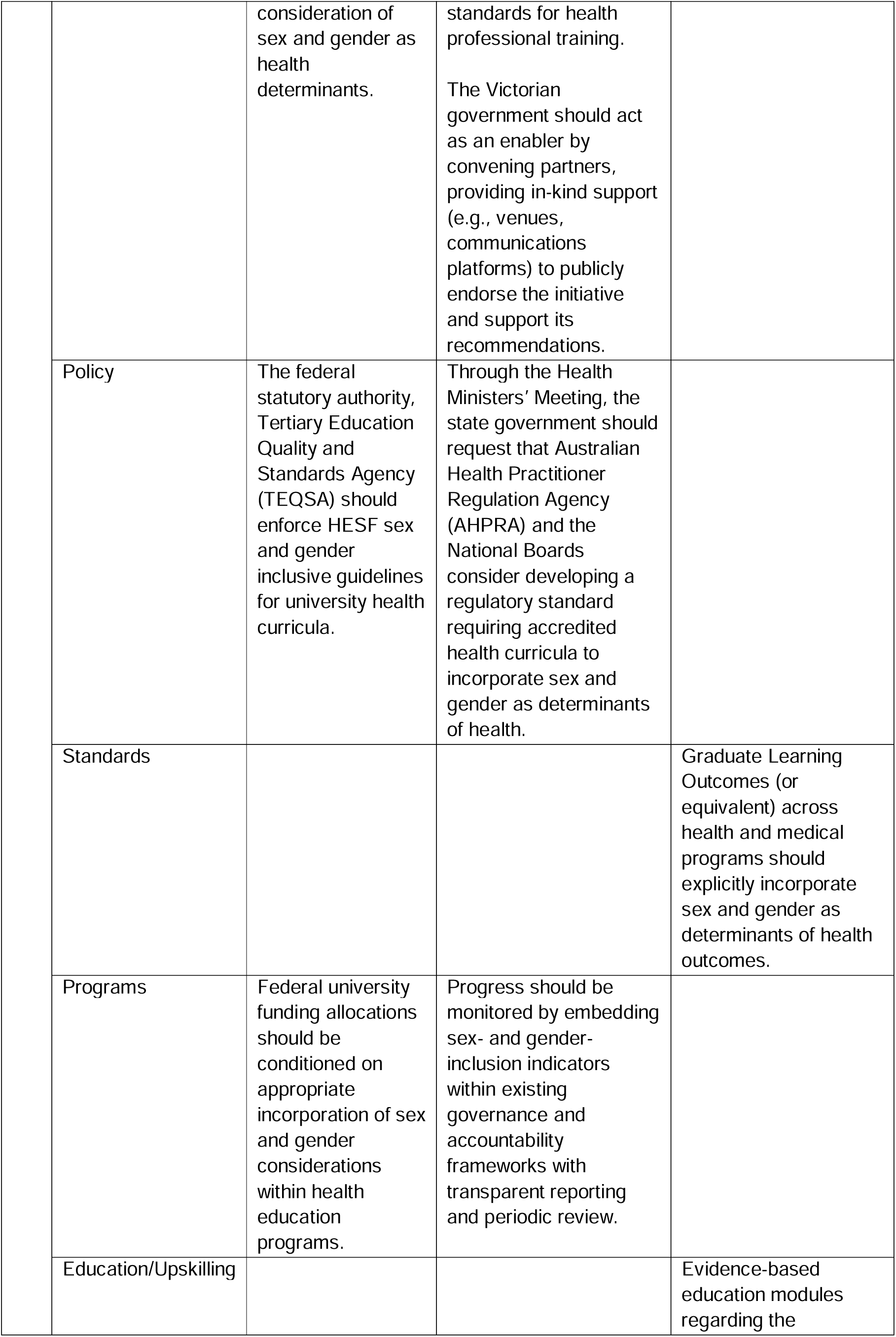
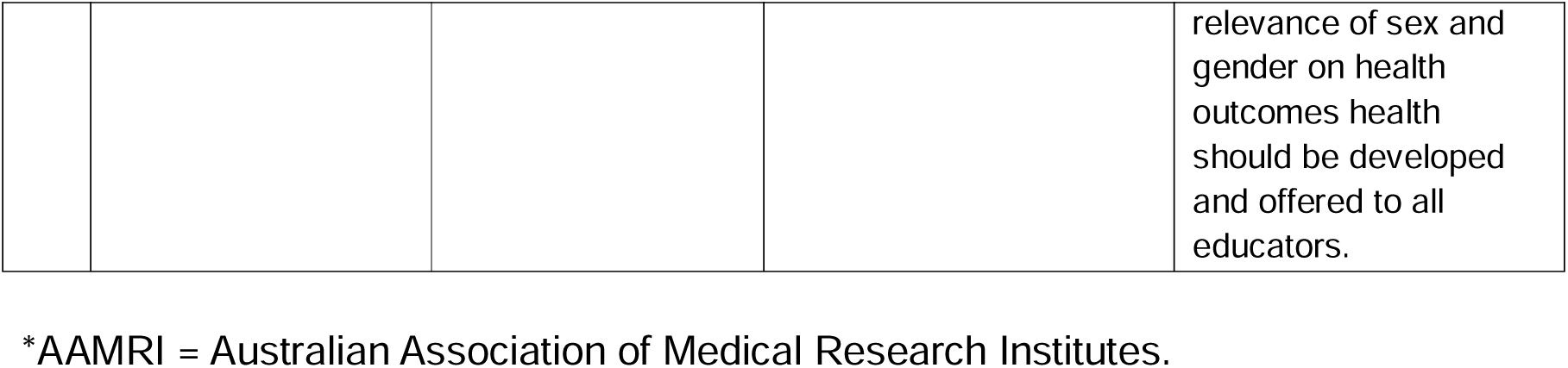
– Recommendations for accelerating inclusion of sex and gender in research and education.

## CONCLUSION

Our findings demonstrate the limited adoption of sex and gender considerations across the Victorian health research and education landscape, serving as a baseline in advance of the new requirement by NHMRC to include sex and gender considerations into funding applications from 2026 [12)]. While this statement represents a significant step forward, it does not obviate the need for other leading research and education bodies across the health and medical sector to develop and implement sex and gender-inclusive policies for the benefit of all Australians. Systematic benchmarking against this robust pre-2026 baseline will reveal the efficacy of implementation and magnitude of impact of such policies, particularly in the current absence of coordinated state-level frameworks.

## Data Availability

All data supporting the findings of this study are available within the manuscript and its supplementary materials. Raw data are available at https://figshare.com/s/b32f49abd615ec38cee5. Supporting information files (S1-S7) are available at: https://doi.org/10.6084/m9.figshare.33093290. Any additional materials, including analytic code are publicly available via the study GitHub repository: https://github.com/severinelamon-prof/Sex-and-gender-incorporation-in-research-in-Victoria/tree/main.

https://doi.org/10.6084/m9.figshare.33093290

https://github.com/severinelamon-prof/Sex-and-gender-incorporation-in-research-in-Victoria/tree/main.

## ACKNOWLEDGMENT PERMISSION

All individuals named as authors have provided their explicit consent to be acknowledged in this article.

## DECLARATIONS

## Acknowledgements

Funding for this work was provided by the Victorian Department of Health, Women’s health and Wellbeing Program to S. Haupt, S. Lamon and R. Huxley, through the Victorian Hub of the Centre of Sex and Gender Equity in Health and Medicine.

## Data sharing statement

All data supporting the findings of this study are available within the manuscript and its supplementary materials. Raw data are available at https://figshare.com/s/b32f49abd615ec38cee5. Supporting information files (S1-S5) are available at: https://doi.org/10.6084/m9.figshare.33093290. Any additional materials, including analytic code are publicly available via the study GitHub repository: https://github.com/severinelamon-prof/Sex-and-gender-incorporation-in-research-in-Victoria/tree/main.

## REFERENCES

1. Heidari S, Babor TF, De Castro P, Tort S and Curno M. Sex and Gender Equity in Research: rationale for the SAGER guidelines and recommended use. Research Integrity and Peer Review. 2016;1(1):2.

2. Haupt S, Carcel C and Norton R. Neglecting sex and gender in research is a public-health risk. Nature. 2024;629(8012):527–530.

3. Flanagan KL and Klein SL. Knowledge gaps and research priorities to understand sex differences in immunity. PLoS Biol. 2026;24(2):e3003578.

4. Mauvais-Jarvis F, Bairey Merz N, Barnes PJ, et al. Sex and gender: modifiers of health, disease, and medicine. Lancet. 2020;396(10250):565–582.

5. Witt A, Norton R, Woodward M and Womersley K. Scientific consideration of sex and gender is the responsibility of the many, not the few. Lancet. 2024;404(10468):2140–2142.

6. Haverfield J and Tannenbaum C. A 10-year longitudinal evaluation of science policy interventions to promote sex and gender in health research. Health Res Policy Syst. 2021;19(1):94.

7. Wainer Z, Carcel C and Sex, Gender Sensitive Research Call to Action Group. Sex and gender in health research: updating policy to reflect evidence. Med J Aust. 2020;212(2):57–62.e1.

8. Haupt S, Carcel C, Halliday L, et al. Catalysing change in health and medical research policy: an Australian case study of deliberative democracy to reform sex and gender policy recommendations. Front Public Health. 2024;12:1522213.

9. Ahmed SB, Becker JB, Bruno KA, et al. Twenty years of sex and gender science in health research: a policy statement from the Organization for the Study of Sex Differences. Biol Sex Differ. 2026;17(1):85.

10. Morrison T, Dinno A and Salmon T. The Erasure of Intersex, Transgender, Nonbinary, and Agender Experiences Through Misuse of Sex and Gender in Health Research. Am J Epidemiol. 2021;190(12):2712–2717.

11. National Health and Medical Research Council. Statement on Sex, Gender, Variations of Sex Characteristics and Sexual Orientation in Health and Medical Research. Canberra: Commonwealth of Australia; 2024. Available from: https://www.nhmrc.gov.au/research-policy/gender-equity/statement-sex-and-gender-health-and-medical-research. Accessed August 2026.

12. National Health and Medical Research Council. Embedding sex and gender considerations in research for better medical outcomes. Canberra: National Health and Medical Research Council; 20 November 2025. Available from: https://www.nhmrc.gov.au/about-us/news-centre/embedding-sex-and-gender-considerations-research-better-medical-outcomes. Accessed August 2026.

13. Carcel C, Vassallo A, Hallam L, et al. Policies on the collection, analysis, and reporting of sex and gender in Australian health and medical research: a mixed methods study. Med J Aust. 2024;221(7):374–380.

14. Merone L, Tsey K, Russell D and Nagle C. Representation of Women and Women’s Health in Australian Medical School Course Outlines, Curriculum Requirements, and Selected Core Clinical Textbooks. Womens Health Rep (New Rochelle). 2024;5(1):276–285.

15. Kirkman M, Honda T, McDonald SJ, et al. Consideration of sex and gender: an analysis of Australian clinical guidelines. Med J Aust. 2025;222(4):205– 209.

16. Baker S. Research funding in Australia: what are the latest trends? Nature Index. 19 December 2024. Available from: https://www.nature.com/nature-index/news/research-funding-australia-latest-trends. Accessed August 2026.

17. Australian Bureau of Statistics. National, state and territory population, June 2025. Canberra: Australian Bureau of Statistics; 2025. Available from: https://www.abs.gov.au/statistics/people/population/national-state-and-territory-population/jun-2025. Accessed August 2026.

18. Eysenbach G. Improving the quality of Web surveys: the Checklist for Reporting Results of Internet E-Surveys (CHERRIES). J Med Internet Res. 2004;6(3):e34.

19. RMIT University. RMIT launches sex-specific pharmacy curriculum to address gender disparities in health. Melbourne: RMIT University; 11 May 2026. Available from: https://www.rmit.edu.au/news/all-news/2026/may/pharmacy-curriculum-addresses-gender-disparities. Accessed August 2026.

20. Bretherton I, Grossmann M, Leemaqz SY, Zajac JD and Cheung AS. Australian endocrinologists need more training in transgender health: A national survey. Clin Endocrinol (Oxf). 2020;92(3):247–257.

21. Zwickl S and Cheung AS. Standardising Gender and Sex Data Collection in Clinical Care and Research. Med J Aust. 2026;224(4):e70177.

22. Seidler ZE, Sheldrake M, Benakovic R, et al. What Does Men’s Health Education Look Like in Australian University Health Curricula? A Formative Evaluation and Future Enhancement Opportunities. J Med Educ Curric Dev. 2024;11:23821205241271564.

23. Njei B, Al-Ajlouni YA, Sidney Kanmounye U, et al. Artificial intelligence agents in healthcare research: A scoping review. PLoS One. 2026;21(2):e0342182.

24. Graham BM. Sex- and gender-responsive management of anxiety disorders: future pathways for research, education, policy and practice. Med J Aust. 2025;223(7):372–378.

25. Gadsden T, Hallam L, Carcel C, et al. Theory of change for addressing sex and gender bias, invisibility and exclusion in Australian health and medical research, policy and practice. Health Res Policy Syst. 2024;22(1):86.

26. White J, Tannenbaum C, Klinge I, Schiebinger L and Clayton J. The Integration of Sex and Gender Considerations Into Biomedical Research: Lessons From International Funding Agencies. J Clin Endocrinol Metab. 2021;106(10):3034–3048.

